# Pregnant Women with Multiple Long-term Conditions Involving Mental Health in Wales, UK (2000-2019): Sequence Clustering Approach in SQL Server Data Mining Using SAIL Databank

**DOI:** 10.1101/2025.03.30.25324916

**Authors:** Mohamed Mhereeg, Siang Ing Lee, Jonathan Kennedy, Muhammad Usman, Lisa Kent, Amaya Azcoaga-Lorenzo, Mairead Black, Colin McCowan, Holly Hope, Kathryn Abel, Krishnarajah Nirantharakumar, Sinead Brophy

## Abstract

**Background:** One in five pregnant women in the UK has multiple long-term conditions (MLTCs), 70% of whom have a mental health condition. MLTCs and mental disorders are independently associated with adverse pregnancy outcomes. Little is known about the sequence in which these conditions develop. Identifying patterns in the accumulation of health conditions can provide valuable insights into disease progression, and potential interaction between conditions, ultimately enhancing our understanding of the complexity of MLTCs in pregnancy.

**Methods:** This retrospective cohort study utilised routinely collected data from the Welsh Secure Anonymised Information Linkage (SAIL) Databank. Eligible women were aged 15 to 49, gave birth in Wales, with a pregnancy start date between 1/1/2000 and 31/12/2019; had pre-existing MLTCs, with at least one mental health condition. MLTCs were defined as having ≥ two out of a predefined list of 22 long-term physical or mental health conditions. The sequence clustering combined sequence analysis (first-order Markov chains) with clustering of sequence of conditions (using Expectation Maximisation) and maternal factors including age, socioeconomic status, BMI and ethnicity.

**Results:** The population cohort included 39,679 women. The most prevalent physical conditions were atopic eczema (33.51%), asthma (33.38%), and allergic rhinoconjunctivitis (30.75%). The most prevalent mental health conditions were depression (27.09%), anxiety (23.01%). The algorithm allocated 35,711 women into 11 sequence clusters. The three most common sequences in the entire cohort were: (i) asthma, depression, other mental health conditions (n=6235, 17.46%); (ii) atopic eczema, depression, other mental health conditions (n=4581, 13%); (iii) other mental health conditions, depression, mixed anxiety and depression (n=3914, 11%). The 11 sequence clusters were predominantly driven by (i) the ordering and (ii) the frequency of the second and third occurring health condition. Within the largest sequence cluster (mental health disorders with subsequent atopic eczema, n=6434, 18.02%), the three most common sequences were: (i) mixed anxiety and depression, depression, atopic eczema; (ii) other mental health conditions, depression, mixed anxiety and depression; (iii) anxiety, depression, atopic eczema. Demographic differences were observed within a few clusters. Younger women aged < 25 (n=4560) were more likely to experience sequences that started with atopic conditions, followed by other mental health conditions and subsequently depression. Women with a BMI over 30 kg/m2 (n=534) were more likely to experience sequences involving conditions such as atopic conditions followed by asthma and depression. A cluster of women with a BMI under 18.5 kg/m² (n=264), from most deprived areas and predominantly White, often started with substance misuse or eating disorders, followed by other mental health conditions and depression.

**Conclusions:** The sequence in which conditions accumulate in this cohort of women with multiple long-term conditions, including mental health disorders, is primarily dominated by mental health and atopic conditions. Women with pre-existing mental illness tend to accumulate both atopic and other mental health conditions throughout their lifetime, contributing to the development of multiple long-term conditions.

## Introduction

One in five pregnant women in the UK starts her pregnancy with multiple long-term conditions (MLTCs) [1]. MLTCs have been associated with adverse maternal and child outcomes [2], [3]. Mental illnesses (primarily mood disorders) are by far the most prevalent conditions in pregnant women with MLTCs and, in recent years, there has been much focus on general discussion on women’s mental health and wellbeing and past mental health history at their first contact with pregnancy services [4]. Moreover, pregnant women with MLTCs accumulate health conditions over their lifetime and the MBBRACE 2023 national maternal mortality review took a step further and recommended the need to target support for vulnerable women with medical and mental health co-morbidities and social complexity [5]

Studying the sequence and clusters of health conditions and associated risk factors may provide better information on MLTC progression over a woman’s lifetime Two recent studies assessing clustering of conditions in pregnancy reported clusters driven by mental health conditions, given its prevalence in pregnant women [6], [7]. However, no study to date considered the temporal order of conditions in pregnant women with MLTC or the pregnant women’s characteristics associated with the sequence of health conditions. By incorporating temporal factors, this analysis overcomes the limitations of traditional clustering methods, offering a deeper understanding of the dynamic relationships and associations between different health conditions. Traditional methods often fail to capture the complex, time-dependent interactions between conditions, such as the transition probabilities between health conditions or the accumulation of multiple long-term conditions. By adding these more sophisticated analytics, our approach accounts for these transitions and temporal dependencies, enabling a more accurate identification of patient trajectories. Additionally, the method employed enabled the examination of the effects of patient characteristics such as age, BMI, ethnicity, and socioeconomic status on cluster formation, providing deeper insight into how these factors influence health condition trajectories.

Therefore, we aimed to examine the sequence of accumulation of health conditions in pregnant women with existing mental health conditions longitudinally in a large, high quality, contemporary UK database. Specifically, we examined health conditions in women prior to pregnancy that emerged before and after mental health conditions using the SAIL national database in Wales.

## Materials and methods

### Study design and inclusion criteria

This is a retrospective cohort study utilising routine health records. Women were eligible if they were aged 15 to 49 at the start of the pregnancy, had a pregnancy start date between 1st January 2000 and 31st December 2019 that resulted in live birth in Wales. They also needed to have multiple long-term conditions (two or more) before the index pregnancy; of which one must be a mental health condition. We selected the last pregnancy in cases where women had multiple recorded pregnancies. Women whose data did not meet standard quality checks were excluded (S1 File).

### Data source

The study utilised anonymised routine health records from the Secure Anonymised Information Linkage (SAIL) for Wales [8]. It is a repository of anonymised health and socio-economic administrative data with population coverage in Wales and provides linkage at an individual level. It holds data for 4.8 million people, contains 100% data coverage from secondary care and has data from and covers 80% of Welsh GP practices [8] Within SAIL, the National Community Child Health Dataset (NCCHD) was employed to identify pregnancies. It was linked to the Welsh Longitudinal General Practice (WLGP) dataset, hospital admissions from the Patient Episode Database for Wales (PEDW), and the Welsh Demographic Service (WDS) dataset for diagnoses, prescriptions, and demographics data.

### Defining maternal multiple long-term conditions

Multiple long-term conditions were defined as the presence of two or more of the 22 pre-existing long-term conditions, with at least one mental health condition, as listed in Box 1 and ranked by their prevalence in the cohort. The selection of 22 conditions was based on a combination of clinical relevance, prevalence in the study cohort, and feasibility for sequence analysis. Including too many conditions could lead to sparsely populated sequences, reducing interpretability and statistical reliability. These conditions were identified using Read Codes and International Classification of Disease-version 10 (ICD10) codes, extracted from primary care records and hospital admissions, respectively. These codes can be found at https://github.com/MMhereeg/MumPredict-Sequence-Clustering. Only the first occurrence of each health condition, prior to the start of pregnancy, was considered in determining the sequence of conditions. While the algorithm identifies and ranks all potential subsequent conditions based on their probability, recurrence was not considered in the sequence identification process in this analysis

**Box 1: 22 long-term conditions used to define maternal multiple long-term conditions**

Mental health conditions

1. Depression
2. Anxiety
3. Other mental health conditions (comprises of obsessive-compulsive disorder, selfharm, personality disorder or dissociative disorder)
4. Mixed anxiety and depression (counted as one condition if both conditions had the same diagnosis dates)
5. Substance misuse
6. Eating disorder
7. Severe mental health conditions (comprises of Bipolar affective disorder, Schizophrenia, or Psychosis).
8. Alcohol misuse/dependency

Physical health conditions

9. Asthma
10. Atopic eczema
11. Allergic rhinoconjunctivitis
12. Migraine
13. Irritable Bowel Syndrome
14. Psoriasis
15. Other skin conditions (comprises of: Seborrheic dermatitis, Rosacea, Hidradenitis suppurativa, Lichen planus)
16. Polycystic Ovary Syndrome
17. Other headaches (comprises of cluster headache, tension headache),
18. Epilepsy
19. Type I or type II diabetes mellitus
20. Neurodevelopmental conditions (comprises of: Learning disability, Attention deficit, hyperactivity disorder, or Autistic spectrum condition)
21. Hypertension
22. Diabetic eye disease

### Sequence clustering method

The sequence clustering employed in this study is a hybrid algorithm that combines sequence analysis with clustering. It is a data mining-based approach that aims to understand the dynamic relationships and associations among health conditions over time [9], [10]. The algorithm uses Markov chain analysis to identify similarly ordered sequences (all possible sequences exist in the data), examines sequence and state transition probabilities, and then combines the results of this analysis with clustering techniques [11], [12]. The algorithm employs the Expectation Maximization algorithm [13] to generate clusters from these sequences based on probability and other variables in the model, including age, BMI, ethnicity and socioeconomic status. This enhancement enabled us to observe variations in clusters and sequences across these variables. The algorithm takes the sequences as inputs and group similar sequences together into clusters. These clusters can then be used to track how patients progress from one condition to another over time.

Additionally, the algorithm includes a Transition Probability Visualization feature, illustrating how individuals progress from one condition to another. As a probabilistic technique, it generates paths that are rarely followed or occur by chance. By incorporating temporal factors, this analysis overcomes the limitations of traditional clustering methods, offering a deeper understanding of the dynamic relationships and associations between different health conditions.

The algorithm requires the clinical diagnoses to be presented as sequence information, stored as a nested table. The data were split into two tables prior to running the algorithm. The first table, the case table, contains the patient ID and demographic information about the patients (non-sequential attributes; age, ethnicity, BMI, and deprivation level). The second table is a nested table that includes a sequence ID column, which identifies the cases within a time sequence, along with the conditions (states) and their corresponding times in the sequence.

This algorithm employed in this study uses first-order Markov chains (Box 2). Each cluster is associated with a first order Markov chain. The number of orders in a Markov chain indicates how many prior states are used to determine the probability of the current state. While higher-order models account for multiple prior states, they increase complexity and introduce computational challenges. A first-order model balances interpretability and computational feasibility while capturing disease progression patterns. In a first-order Markov model, the probability of the current state depends solely on the previous state. A Markov chain is characterized by a finite set of N allowed states, denoted as S = {S1, S2, …, Sn}, and the Markov property, which means that the state at time t, denoted by s(t), depends only on the preceding state s(t−1) and not on earlier states such as s(t−2), s(t−3), etc. This property is expressed by means of transition probabilities in our context, the state-space S consists of the various conditions recorded in the patient’s medical record.

**Box 2: How the algorithm works**

- The algorithm takes as input parameter the number *K* of clusters
- Assumes an initial set of clusters C = {C1, C2, …, Ck}, this can be adjusted and defined manually.
- Each cluster *c_k_* is a Markov chain on its own (NxN transition matrix).
- The algorithm can be described as follows:

1. Initialises randomly the transition matrix of each cluster.
2. Looks for sequences and examines all transition probabilities and measures the differences between all the possible sequences in the dataset
3. Determines which sequences are the best candidates to use as inputs for clustering.
4. **Expectation step**—assign each sequence to the cluster that can produce it with higher probability
5. **Maximisation step**—recompute the transition matrices for all clusters by considering the set of sequences assigned to each cluster in the previous step.
6. **Repeat steps 4 and 5** iteratively until the transition matrices do not change; at this point, the assignment of sequences to clusters also does not change anymore.
The result is a set of Markov models that describes the behaviour of each cluster.

**Figure 1:**
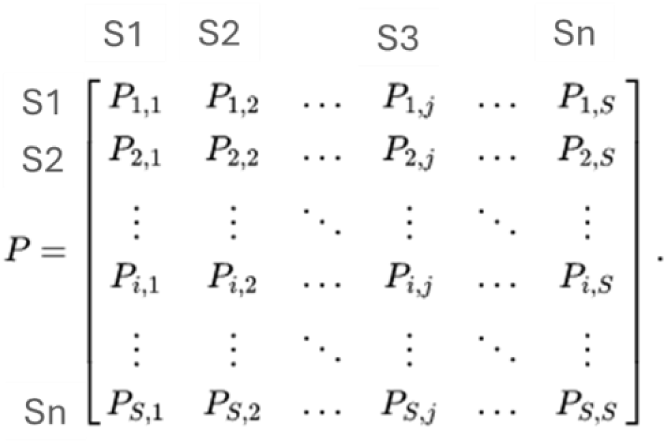
Markov chain transition matrix, each state S1, S2, …, Sn represents a possible condition in a patient’s medical record. The transition probability P between two states (e.g, from Si to Sj) reflects the likelihood of moving from one condition to another, based on the previous state.

### Optimising performance and accuracy of the mining model

The Microsoft Sequence Clustering algorithm includes parameters that influence the behaviour, performance, and accuracy of the resulting mining model. These parameters include Cluster Count (which defines the approximate number of clusters to be built by the algorithm), Minimum Support (which sets the minimum number of cases required to form a cluster), and Maximum_Sequence_States (which specifies the maximum number of states a sequence can have).

Initially, we used the default settings, which resulted in 17 clusters. However, this high number of clusters made the analysis rather difficult to interpret and analyse. After experimenting with different values as recommended, we found that setting the Cluster Count to 10, Minimum Support to 1000, and Maximum_Sequence_States to 64 yielded more intuitive results that were easier to analyse. Generally, too many clusters can complicate the identification of typical behaviours, as even small variations can cause similar patterns to be spread across multiple sequence clusters with low support. Conversely, using too few clusters can combine distinct behaviours into a single group, resulting in overly complex models that are difficult to interpret and analyse.

## Results

### Study population

Out of a total 587,718 pregnancies in the Welsh population during the study period, 49,758 (8.47%) pregnancies resulted in live births had sequences of maternal health conditions with at least one mental health condition were included in the analysis (Fig. 2, describes the flowchart of participants selection). 80% of data (n=39,679 pregnancies) were used to train the model and will be considered as the study cohort; 20% (n=10,079 pregnancies) were reserved for testing model accuracy. The main findings from the 39,679 pregnancies are presented in Table 1 (characteristics of the 39,679 included pregnancies). Most pregnancies (n= 32,503 pregnancies) were in women aged 20 -35 years at conception (81.92%); 60.61% were of White ethnicity; 43.92% were overweight or obese (BMI ≥25kg/m^2^, and 27.56% were in the most deprived quintile. The median number of conditions recorded prior to index pregnancy was 3 (IQR 2-4). Most prevalent mental health conditions evident prior to conception were depression (27.09%), anxiety (23.01%) and other mental health conditions (18.97%). The most prevalent accompanying physical health conditions were atopic eczema (33.51%), asthma (33.38%), and allergic rhinoconjunctivitis (30.75%).

**Fig 2.**
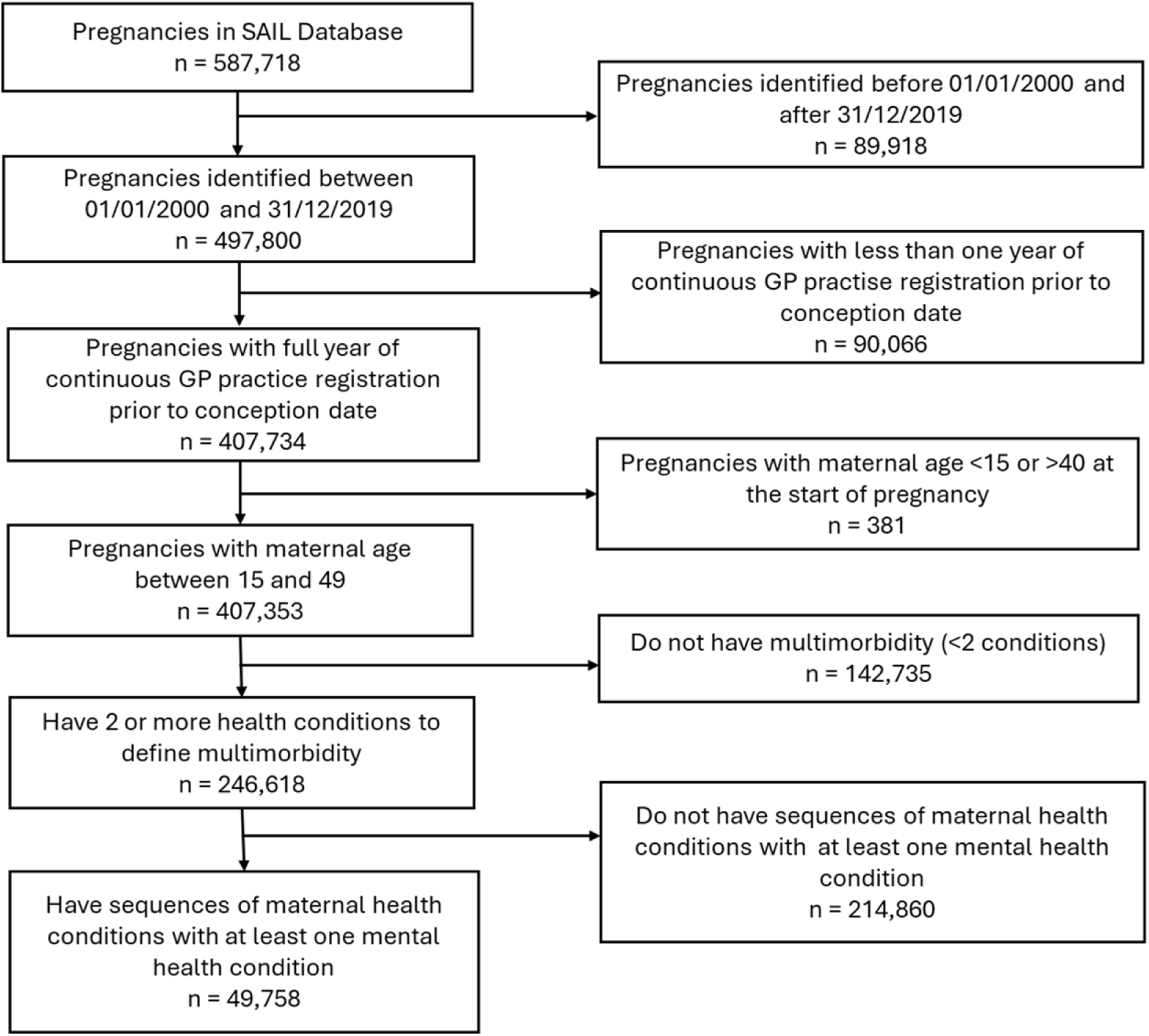
Flowchart of participant selection.

**Table 1:** Characteristics of the women of the 39,769 included pregnancies.

| Characteristics | N (%) |
| --- | --- |
| **Cluster size** | 39,679 |
| **Number of conditions** |  |
| Median (Interquartile range) | 3 (2 to 4) |
| Mean (Standard deviation) | 3 (1.33) |
| **Maternal age at conception, years** |  |
| 15-19 | 2646 (6.67%) |
| 20-24 | 8884 (22.39%) |
| 25-29 | 11792 (29.72%) |
| 30-35 | 11827 (29.81%) |
| 36-49 | 4530 (11.42%) |
| **Ethnicity** |  |
| Asian | 200 (0.5%) |
| Black | 134 (0.34%) |
| Mixed | 104 (0.26%) |
| Other | 101 (0.25%) |
| White | 24050 (60.61%) |
| Missing | 15090 (38.03%) |
| **Body mass index, kg/m<sup>2</sup>** |  |
| <18.5 | 2210 (5.57%) |
| 18.5-24.9 | 15280 (38.51%) |
| 25-29.9 | 8859 (22.33%) |
| 30+ | 8566 (21.59%) |
| Missing | 4764 (12.01%) |
| **Welsh Index of Multiple Deprivation, quintile** |  |
| 1, most deprived | 10936 (27.56%) |
| 2 | 8437 (21.26%) |
| 3 | 6840 (17.24%) |
| 4 | 5595 (14.1%) |
| 5, least deprived | 5325 (13.42%) |
| Missing | 2546 (6.42%) |
| **Health conditions pre-conception** |  |
| Atopic eczema | 13296 (33.51%) |
| Asthma | 13246 (33.38%) |
| Allergic Rhinoconjunctivitis | 12201 (30.75%) |
| Depression | 10750 (27.09%) |
| Anxiety | 9130 (23.01%) |
| Migraine | 8642 (21.78%) |
| Other mental health conditions (self-harm/suicide, obsessive compulsive disorder, personality disorder, dissociative disorder) | 7525 (18.97%) |
| Irritable Bowel Syndrome | 5963 (15.03%) |
| Mixed anxiety and depression | 5409 (13.63%) |
| Other skin conditions (Seborrheic dermatitis, Rosacea, Hidradenitis suppurativa, Lichen planus) | 4050 (10.21%) |
| Other headache (tension headache, cluster headache, other chronic headache) | 2884 (7.27%) |
| Polycystic ovarian syndrome | 2674 (6.74%) |
| Psoriasis | 2351 (5.92%) |
| Alcohol misuse/dependency | 1933 (4.87%) |
| Eating disorder | 1483 (3.74%) |
| Substance misuse/dependency | 1400 (3.53%) |
| Epilepsy | 864 (2.18%) |
| Hypertension | 693 (1.75%) |
| Diabetes mellitus (type I and II) | 601 (1.51%) |
| Severe mental illness (bipolar disorder / affective psychosis, Schizophrenia / non-affective psychosis) | 535 (1.35%) |
| Neurodevelopmental conditions | 394 (0.99%) |
| Diabetic eye disease | 240 (0.60%) |

### Sequence clustering analysis of health conditions

In total, 11 sequence clusters were generated (Table 2). There were 35,711 (90%) of 39,679 pregnancies in the study cohort allocated to these 11 clusters. The remaining 3,968 women were not allocated to any of the clusters because the Markov transition matrix stores only sequences with a probability greater than 0. Figure 1a shows how these clusters are associated, with similar clusters grouped close together. The shade of each node represents the density of all cases in the cluster: the darker the shade, the more cases it contains. The edges represent the similarity between clusters and how clusters are associated, thicker edges mean higher similarity. The algorithm assigns women to clusters taking into account any of the 22 conditions they have as well as their age, BMI, Ethnicity, and deprivation level. Figures 1b, 1c. and 1d show clusters generated for younger women aged 15-19, those with a BMI 30 kg/m2 or over, and those from most deprived areas respectively. Women aged 15-19 are mainly assigned by the algorithm to the [Atopic conditions followed by mental health disorders and subsequently depression] cluster and those with a BMI over 30 kg/m2 were assigned to the [Atopic conditions followed by asthma and depression] and [polycystic ovarian syndrome or skin conditions followed by depression] clusters and the other relatively dark clusters Involving atopic and mental health conditions. Women from the most deprived areas were primarily assigned to clusters associated with eating disorders, substance misuse, atopic conditions, and mental health disorders. These include clusters such as [Eating disorders or substance misuse followed by other mental health conditions and depression], [Atopic conditions followed by mental health disorders and subsequently depression], and [Other mental health or atopic conditions followed by depression and mixed anxiety and depression], as well as other relatively dark clusters involving migraine, asthma, and skin conditions (S2A-S2D Fig. Clusters revealed in the data within subgroups, presents the clusters revealed in the data by age, BMI, deprivation level, and ethnicity subgroups).

**Table 2:** characteristics of the sequence clusters.

| Cluster No | Cluster name | Cluster size |  | Number of conditions |  |
| --- | --- | --- | --- | --- | --- |
|  |  | N | % | Median (IQR) | Mean (SD) |
| 1 | Mental health disorders with subsequent atopic eczema | 6434 | 18.02 | 2 (2 to 3) | 2.50 (0.82) |
| 2 | Atopic conditions followed by mental health disorders and subsequently depression | 5629 | 15.76 | 3 (2 to 4) | 3.04 (1.35) |
| 3 | Allergies or migraine with subsequent anxiety and depression | 4429 | 12.40 | 3 (2 to 3) | 2.85 (1.11) |
| 4 | Atopic conditions followed by migraine and anxiety | 3946 | 11.05 | 3 (2 to 4) | 3.18 (1.20) |
| 5 | Allergies, IBS or asthma with atopic eczema and anxiety | 3092 | 8.66 | 3 (2 to 4) | 3.05 (1.15) |
| 6 | Atopic conditions and migraine followed by asthma and depression | 2742 | 7.68 | 3 (2 to 4) | 3.14 (1.29) |
| 7 | Other mental health or atopic conditions followed by depression and mixed anxiety and depression | 2426 | 6.79 | 2 (2 to 3) | 2.78 (1.06) |
| 8 | Other mental health or atopic conditions followed by allergies and depression | 2389 | 6.69 | 3 (2 to 4) | 3.37 (1.24) |
| 9 | Eating disorders or substance misuse followed by other mental health conditions and depression | 2143 | 6.00 | 3 (2 to 4) | 3.19 (1.28) |
| 10 | Atopic conditions followed by asthma and depression | 1383 | 3.87 | 4 (3 to 5) | 3.91 (1.30) |
| 11 | Polycystic ovarian syndrome or skin conditions followed by depression | 1098 | 3.07 | 3 (2 to 4) | 3.37 (1.32) |

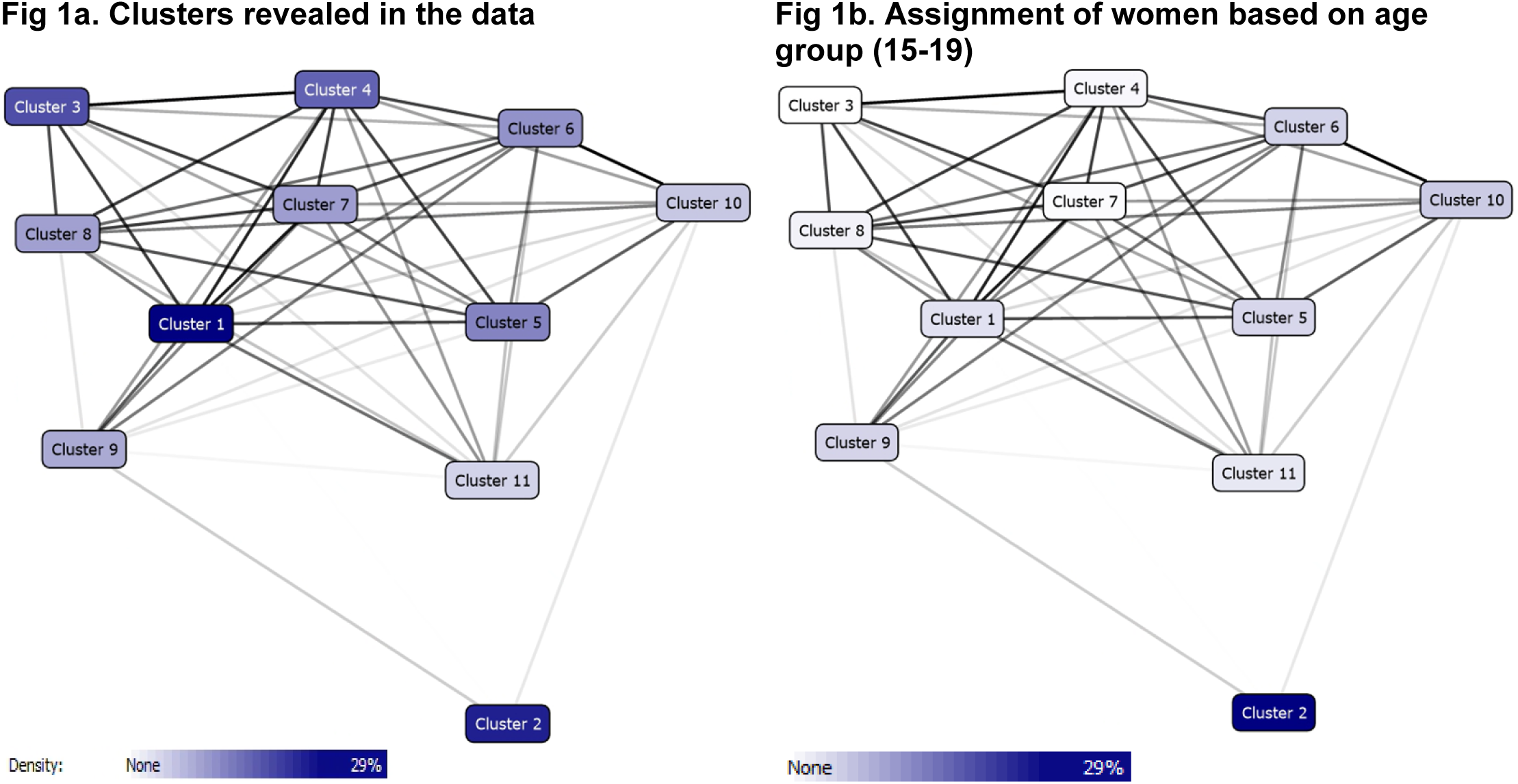
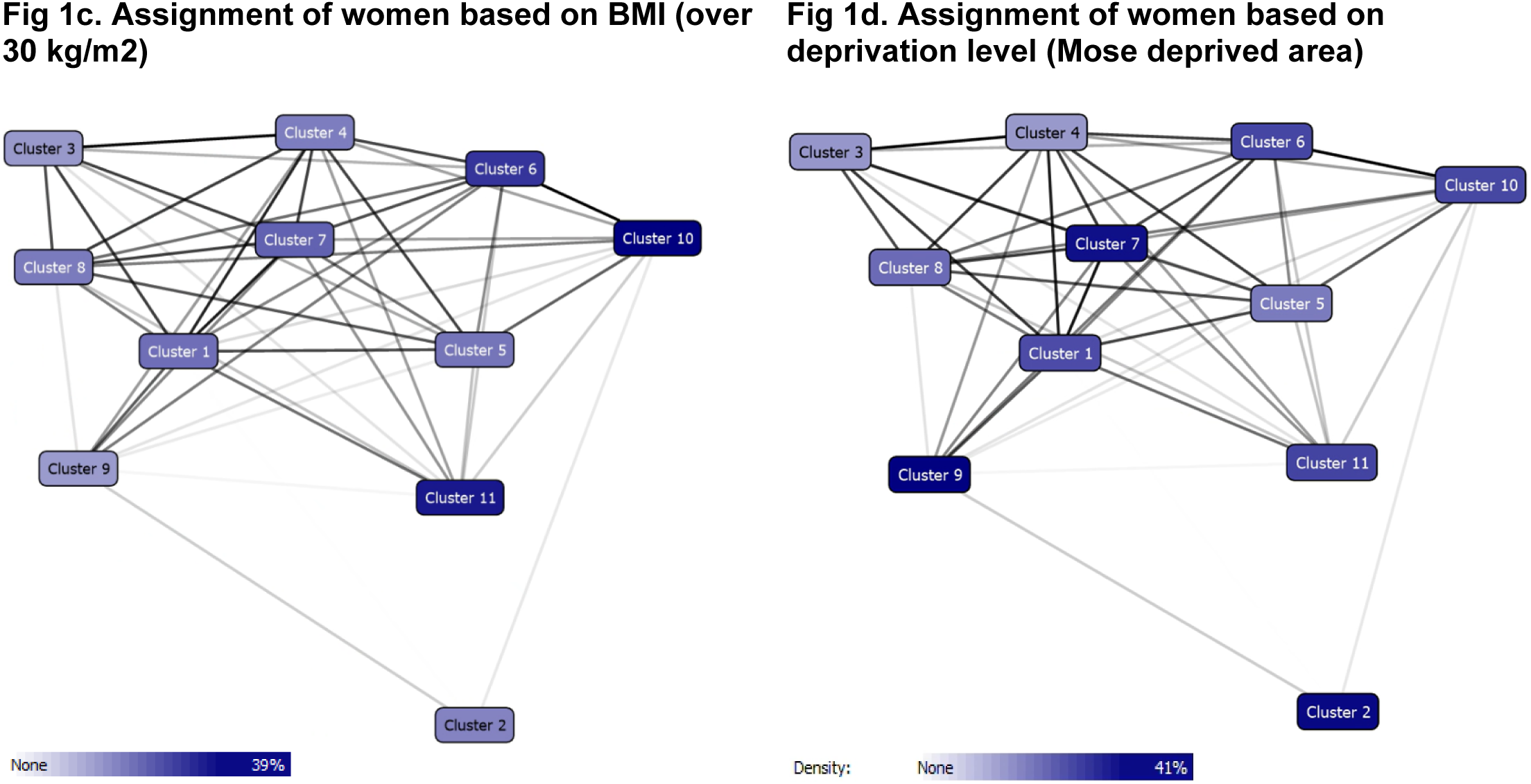

### Most common sequences of health conditions in the study cohort

Table 3 presents the top 10 most common sequences of health conditions in the study cohort. These were driven mainly by health conditions that were most prevalent preconception in the study cohort. Pregnant women in this study cohort (who have at least 1 mental health condition by the study criteria) most commonly started with an atopic condition (atopic eczema, asthma), followed by a mental health condition. Another sequence that was common was an accumulation of different mental health conditions overtime before pregnancy. For example, one of the top 10 most common sequences started with mixed anxiety and depression, followed by depression, then other mental health conditions.

**Table 3:** The top 10 most common sequences of health conditions in the study cohort.

| Sequence size | 1<sup>st</sup> condition | n/N Probability (%) | 2<sup>nd</sup> condition | Probability (%) | 3<sup>rd</sup> condition | Probability (%) |
| --- | --- | --- | --- | --- | --- | --- |
| 6236 | Asthma | 6235/35711 (17%) | Depression | 535/6235 (9%) | Other mental health conditions | 37/535 (7%) |
| 4581 | Atopic eczema | 4581/35711 (13%) | Depression | 427/4581 (9%) | Other mental health conditions | 29/427 (7%) |
| 3914 | Other mental health conditions | 3914/35711 (11%) | Depression | 555/3914 (14%) | Mixed anxiety & depression | 38/555 (7%) |
| 3780 | Mixed anxiety & depression | 3780/35711 (11%) | Depression | 341/3780 (9%) | Other mental health conditions | 23/341 (7%) |
| 3500 | Depression | 3500/35711 (10%) | Other mental health conditions | 241/3500 (7%) | Mixed anxiety & depression | 34/241 (6%) |
| 3363 | Allergic rhinoconjunctivitis | 3363/35711 (9%) | Asthma | 474/3363 (14%) | Depression | 41/474 (9%) |
| 2828 | Anxiety | 2828/35711 (8%) | Mixed anxiety & depression | 203/2828 (7%) | Depression | 18/203 (9%) |
| 2111 | Migraine | 2111/35711 (6%) | Depression | 235/2111 (11%) | Other mental health conditions | 16/235 (7%) |
| 1084 | Irritable bowel syndrome | 1084/35711 (3%) | Anxiety | 128/1084 (12%) | Mixed anxiety & depression | 9/128 (7%) |
| 781 | Other skin conditions | 781/35711 (2%) | Atopic eczema | 84/781 (11%) | Depression | 8/84 (9%) |

### Most common second condition following the starting condition

Table 4 lists the 22 health conditions included in this study and presents the most common second conditions that developed next. A mental health condition was the most common second condition to develop, following either a physical or mental health first condition. This was also the case for less common mental health conditions, such as eating disorder (most commonly followed by depression), substance misuse/dependency (most commonly followed by other mental health conditions), and severe mental illness (most commonly followed by depression). The exceptions were allergic rhinoconjunctivitis (the most common second condition that followed was asthma), alcohol misuse/dependency (most commonly followed by allergic rhinoconjunctivitis) and diabetic eye disease (most commonly followed by diabetes mellitus). Of note, for type I or II diabetes mellitus, the top 3 most common second conditions were depression, diabetic eye disease and asthma.

**Table 4:**
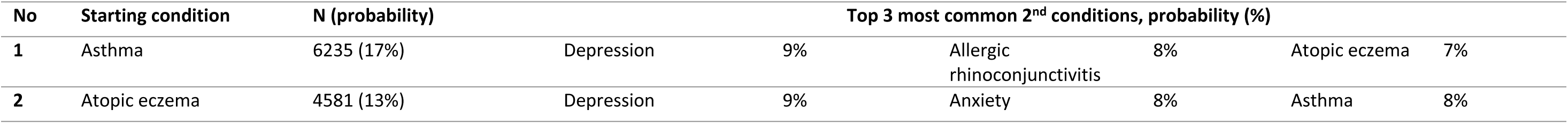
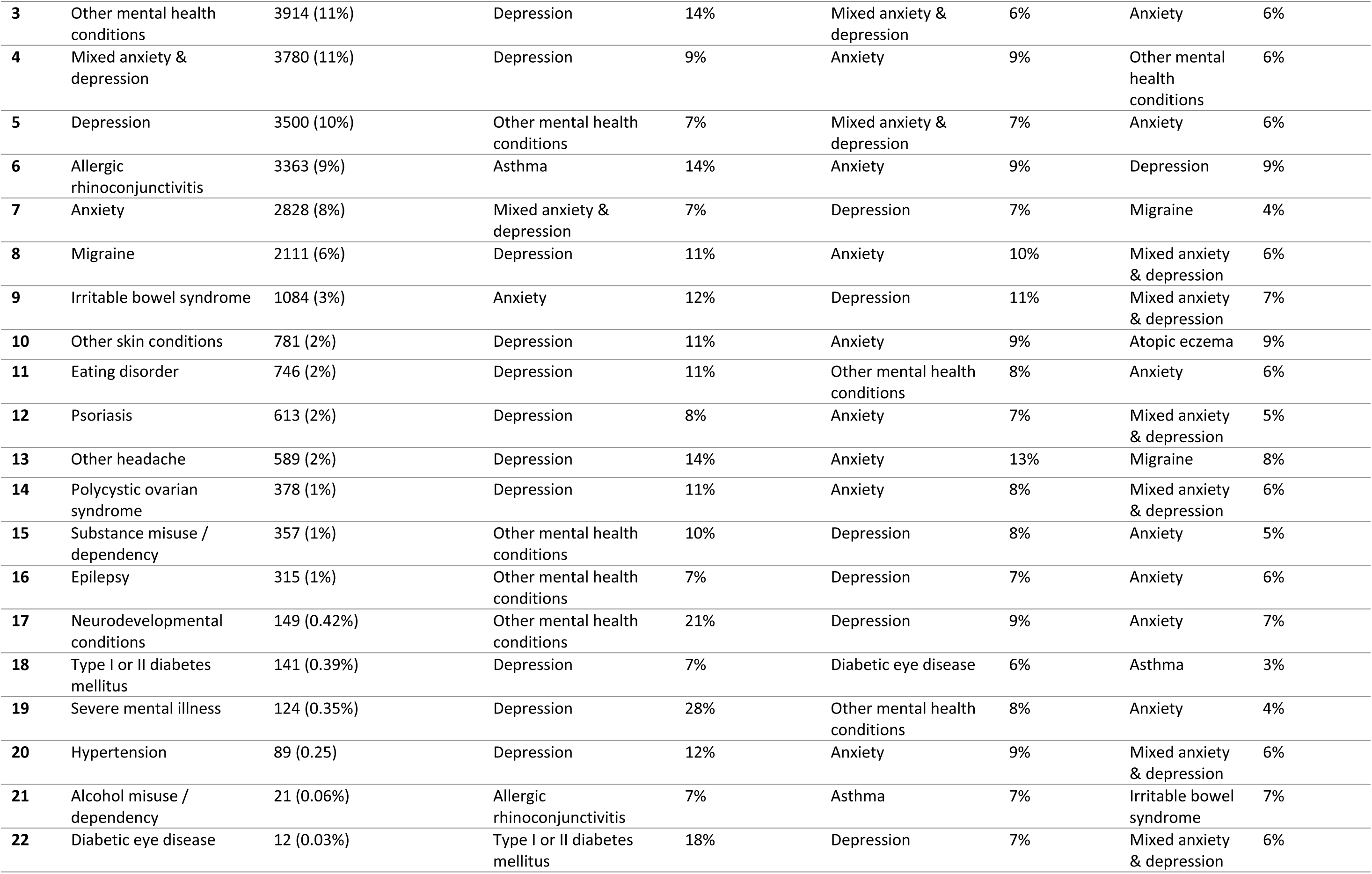
The most common 2^nd^ conditions to develop after the starting condition for the 22 health conditions.

### Most common sequences of health conditions within the 11 sequence clusters

Table 5 presents the top 3 most common sequences of health conditions for the 11 sequence clusters. It presents the 1st starting condition, followed by the most commonly occurring 2nd condition for the listed 1st condition, followed by the most common 3rd condition after the listed 2nd condition. The sequence clusters are predominantly driven by (i) the order and (ii) the frequency of the second and third occurring health conditions within the cluster. For example, in Cluster 2 [Atopic conditions followed by mental disorders] (15.76%), for the top 3 most common sequences, despite having different starting conditions, all had the same second occurring conditions (other mental health conditions, also the most prevalent condition) and the same third condition (depression).

**Table 5:**
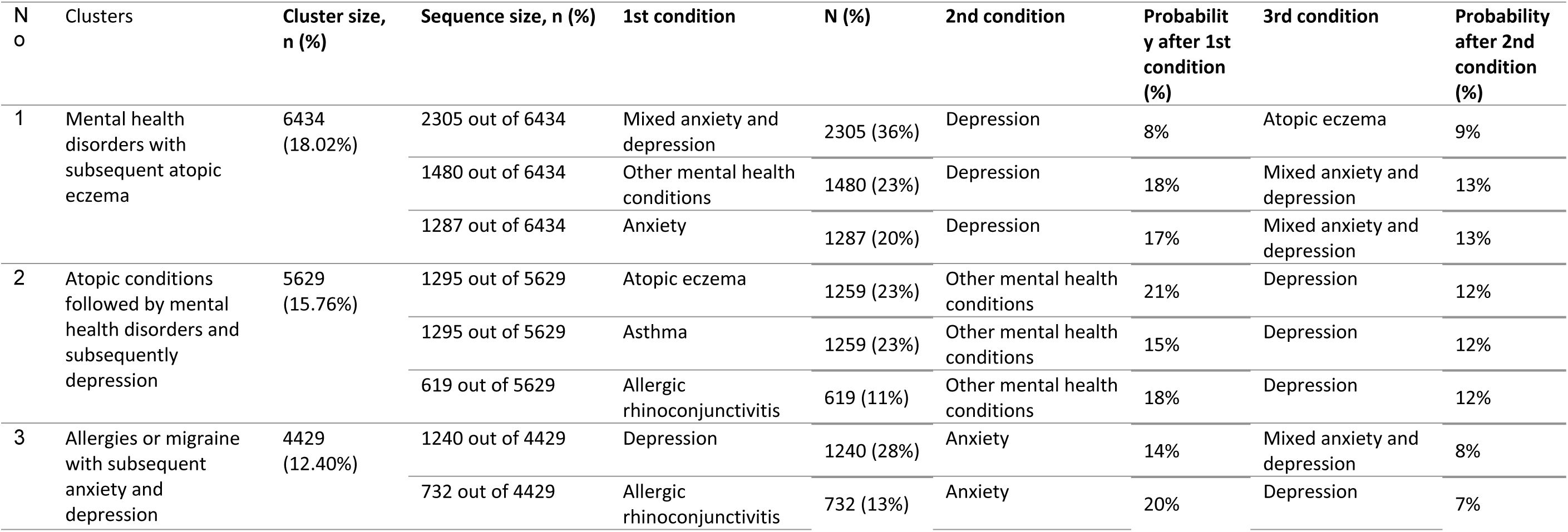
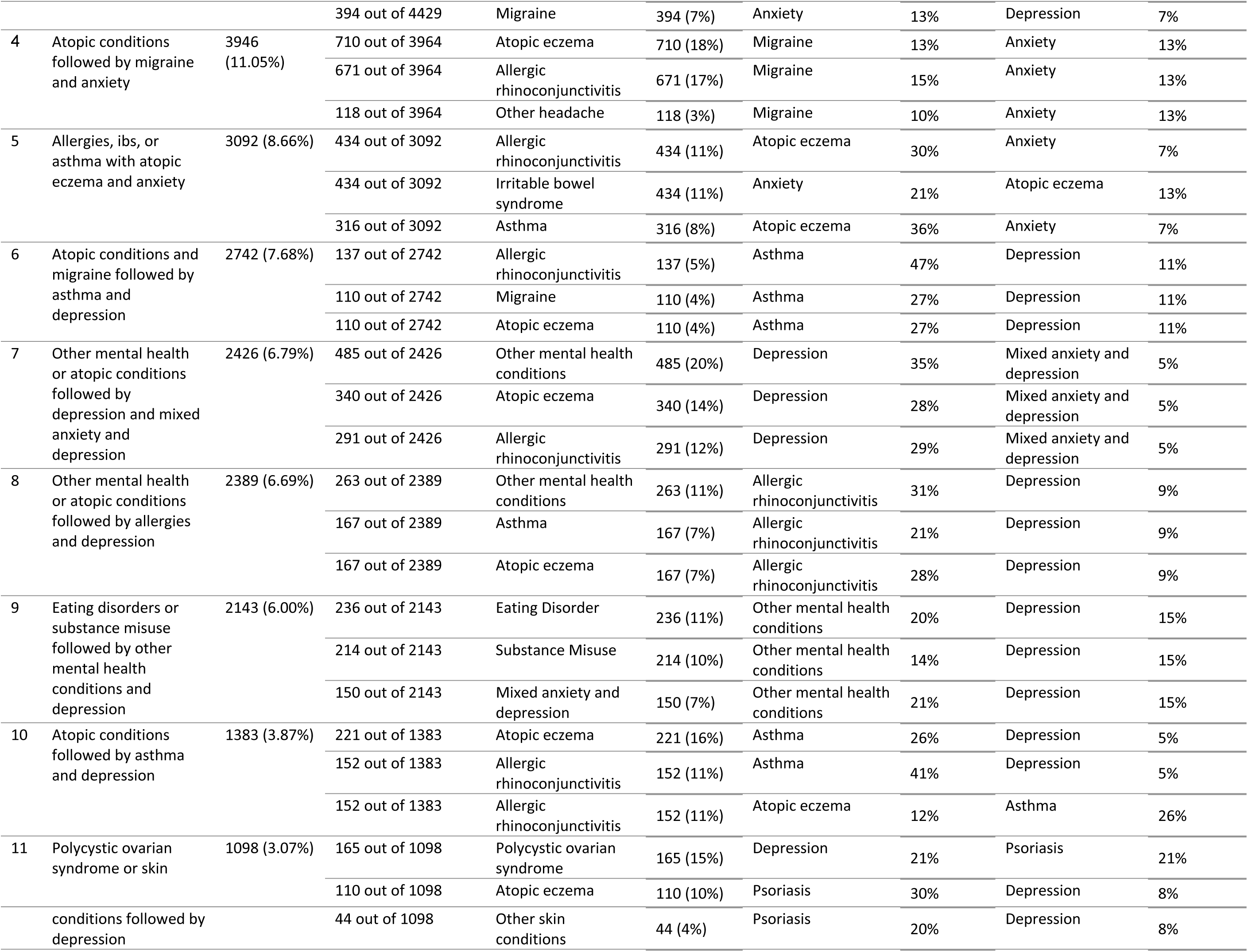
The top 3 most common sequence of health conditions within the 11 sequence clusters.

The top 3 largest clusters all had mental health conditions as their most common second and third order conditions. Most clusters featured sequences with (i) an accumulation of mental health conditions, (ii) an accumulation of atopic conditions (i.e, asthma, atopic eczema, allergic rhinoconjuncitivits), or (iii) started with atopic conditions and subsequently followed by mental health conditions.

However, 3 further clusters showed distinct patterns to these. Cluster 4 [Atopic conditions followed by migraine and anxiety] had sequences that started with atopic conditions, followed by migraine, then anxiety. Cluster 9 [Eating disorders or substance misuse followed by other mental health conditions and depression] started with substance misuse or eating disorder, which had low prevalence in the study cohort (3.5% and 3.7%, respectively). These were then followed by other mental health conditions, and subsequently depression. Cluster 11 [Polycystic ovarian syndrome or skin conditions followed by depression] featured psoriasis being preceded by atopic eczema or other skin condition followed by depression.

### Characteristics of women within the sequence clusters

Table 6 presents the characteristics of the women within the clusters. Here we highlight clusters with demographics that differ from the study cohort and the other clusters.

**Table 6:** Characteristics of the pregnant women by the 11 sequence clusters.

| Characteristics, N (%) | Cluster 1 | Cluster 2 | Cluster 3 | Cluster 4 | Cluster 5 | Cluster 6 | Cluster 7 | Cluster 8 | Cluster 9 | Cluster 10 | Cluster 11 |
| --- | --- | --- | --- | --- | --- | --- | --- | --- | --- | --- | --- |
| **Total** | 6434<br>(18.02%) | 5629<br>(15.76%) | 4429<br>(12.4%) | 3946<br>(11.05%) | 3092<br>(8.66%) | 2742<br>(7.68%) | 2426<br>(6.79%) | 2389<br>(6.69%) | 2143<br>(6%) | 1383<br>(3.87%) | 1098<br>(3.07%) |
| **Maternal age at conception, years** |  |  |  |  |  |  |  |  |  |  |  |
| **15-19** | **3%** | 29% | <0.05 | 1% | 5% | 5% | 0% | 2% | 5% | 6% | 3% |
| **20-24** | **23%** | 52% | 5% | 14% | 23% | 18% | 14% | 12% | 26% | 26% | 16% |
| **25-29** | **32%** | 17% | 24% | 32% | 35% | 31% | 37% | 32% | 32% | 35% | 31% |
| **30-35** | **31%** | 2% | 48% | 39% | 28% | 32% | 35% | 39% | 27% | 26% | 36% |
| **36-49** | **12%** | 0% | 23% | 14% | 9% | 15% | 13% | 15% | 10% | 7% | 15% |
| **Ethnicity** |  |  |  |  |  |  |  |  |  |  |  |
| **Asian** | **1%** | 0% | 1% | 1% | 1% | 0% | 0% | 2% | 0% | 0% | 1% |
| **Black** | **0%** | 0% | 0% | 0% | 0% | 0% | 1% | 1% | 0% | 1% | 1% |
| **Mixed** | **0%** | 0% | 0% | 0% | 0% | 0% | 1% | 1% | 0% | 0% | 0% |
| **Other** | **0%** | 0% | 0% | 0% | 0% | 0% | 0% | 0% | 0% | 0% | 0% |
| **White** | **57%** | 68% | 56% | 64% | 57% | 64% | 44% | 53% | 75% | 67% | 60% |
| **Missing** | **41%** | 31% | 43% | 35% | 42% | 35% | 53% | 44% | 24% | 33% | 38% |
| **Body mass index, kg/m²** |  |  |  |  |  |  |  |  |  |  |  |
| **<18.5** | **5%** | 9% | 3% | 6% | 5% | 5% | 5% | 4% | 12% | 3% | 3% |
| **18.5-24.9** | **34%** | 31% | 51% | 45% | 39% | 35% | 38% | 43% | 45% | 29% | 31% |
| **25-29.9** | **22%** | 17% | 24% | 23% | 25% | 24% | 25% | 27% | 18% | 24% | 24% |
| **30+** | **22%** | 19% | 15% | 20% | 21% | 31% | 24% | 20% | 15% | 39% | 35% |
| **Missing** | **17%** | 25% | 7% | 7% | 11% | 6% | 9% | 6% | 10% | 5% | 7% |
| **Welsh Index of Multiple Deprivation, quintile** |  |  |  |  |  |  |  |  |  |  |  |
| **1, most deprived** | **28%** | 39% | 17% | 16% | 21% | 29% | 38% | 24% | 41% | 29% | 30% |
| **2** | **22%** | 26% | 17% | 21% | 21% | 23% | 19% | 19% | 20% | 19% | 20% |
| **3** | **19%** | 14% | 16% | 20% | 19% | 16% | 12% | 16% | 17% | 20% | 18% |
| **4** | **14%** | 9% | 19% | 18% | 16% | 14% | 12% | 17% | 10% | 12% | 12% |
| **5, least deprived** | **10%** | 5% | 25% | 17% | 16% | 12% | 14% | 19% | 6% | 12% | 14% |
| **Missing** | **6%** | 7% | 7% | 7% | 7% | 6% | 4% | 6% | 7% | 8% | 6% |
| **Top 3 most common health conditions, %** |  |  |  |  |  |  |  |  |  |  |  |
|  | Mixed anxiety & depression (24%) | Other mental health conditions (25%) | Anxiety (30%) | Migraine (17%) | Atopic eczema (44%) | Asthma (43%) | Depression (41%) | Allergic rhinoconjunctivitis (37%) | Other mental health conditions (28%) | Asthma (45%) | Psoriasis (34%) |
|  | Depression (17%) | Asthma (18%) | Depression (17%) | Anxiety (13%) | Anxiety (11%) | Depression (12%) | Mixed anxiety & depression (13%) | Depression (12%) | Depression (16%) | Atopic eczema (14%) | Depression (10%) |
|  | Other mental health conditions (15%) | Depression (11%) | Mixed anxiety & depression (12%) | Depression (11%) | Depression (9%) | Anxiety (7%) | Other mental health conditions (8%) | Anxiety (8%) | Substance misuse (14%) | Allergic rhinoconjunctivitis (12%) | Anxiety (7%) |

Cluster 2 [Atopic conditions followed by mental health disorders] consists predominantly of woman aged under 25 years old (81.40% [95% CI 80.40 - 82.40] compared to 29.06% [95% CI 28.60 - 29.50] of the study cohort); this cluster predominantly starts with an atopic condition, followed by other mental health conditions and subsequently depression.

Cluster 9 [Eating disorders or substance misuse followed by other mental health conditions and depression] had higher proportion of women who were underweight (BMI <18.5 kg/m2, 12.30% [95% CI 10.90 - 13.70] compared to 5.57% [95% CI 5.30 - 5.80] of the study cohort), most deprived (40.60% [95% CI 38.50 - 42.70] versus 27.56% [95% CI 27.10 - 28.00]), and of White ethnicity (75% [95% CI 73.20 - 76.80] versus 60.61% CI [95% 60.10 - 61.10]); this cluster predominantly starts with substance misuse or eating disorder, followed by other mental health conditions and subsequently depression.

Cluster 10 [Atopic conditions followed by asthma and depression] and 11 [Polycystic ovarian syndrome or skin conditions with depression] had higher proportion of women who had BMI >30 kg/m2 (38.60% [95% CI 36.00 - 41.20] and 34.80% [95% CI 32.00 - 37.60], respectively, compared to 21.59% [95% CI 21.20 - 22.00] of the study cohort). Cluster 10 starts with atopic conditions (allergic rhinoconjunctivitis or atopic eczema), followed by asthma and depression. Cluster 11 starts with a range of different conditions, one of which is polycystic ovarian syndrome, followed by psoriasis and depression subsequently.

## Discussion

### Main findings

The most common sequences started with atopic conditions followed by mental health conditions. Following development of a physical or mental health long-term condition, the most common second condition was another mental health condition. The algorithm allocated women into 11 sequence clusters. The sequence clusters were generally an accumulation of mental health conditions, atopic conditions, or started with atopic conditions and subsequently followed by mental health conditions. The defining feature of the sequence clusters was having the same second and third health conditions. Our findings are consistent with previous evidence on pregnant women reporting a positive association between atopic conditions and mental conditions (specifically depression) [14][15] between substance misuse and depression [16]; eating disorder and depression [17]; and between atopic conditions and high body mass index [18][19].

Two of the 11 sequence clusters (clusters 1 and 9) featured 3 mental health conditions, and the other sequence clusters also featured accumulation of mental health conditions. Previous studies [20][21] also demonstrate the risk of subsequent mental ill health is greater among those who develop a mental illness at a younger age (before age 20).

### Findings in the context of existing literature

A Canadian study (Brown et al, 2024) reported that mood and anxiety disorders (77%) were the most prevalent pre-existing long-term conditions in pregnant women with multiple long-term conditions [22]. Brown et al’s cross sectional latest class cluster analysis found the clusters were driven by mental health conditions, with 4 out of 5 clusters featuring mood and anxiety disorder, plus one other health condition. Given the high prevalence of mental illness in young people and in women of reproductive age, perhaps this is unsurprising. The present study, therefore, focused on pregnant women with multiple long-term condition including at least one mental health condition prior to conception and our sequence clusters were also dominated by the most common mental health conditions, anxiety and depression.

Using the UK Biobank, S. Han et al (2024) examined progression of MLTCs for 190 health conditions [23] and reported that anxiety was amongst the top 10 conditions most likely to develop after other health conditions. Common mental health conditions (anxiety and depression) were also frequently observed as second and third order conditions in our sequence analysis. In the Biobank study, for females, the top 10 influential conditions that were most likely to be associated with progression to other conditions additionally included two related to childbirth: labour and delivery complicated by fetal distress [ICD-10 O68] and perineal laceration during delivery [O70]. Although this finding relates to our population of interest, and pregnant women with obstetric complications are at increased risk of accumulating further long-term conditions ahead of their next pregnancy, evidence from UK Biobank is skewed towards a much older, less sociodemographically diverse population. And it is less likely, therefore, to be able to answer questions about the progression of MLTCs in which mental disorders predominate Moreover, a study utilized a predefined list of 77 long-term conditions and routine health records from two separate UK databases, the Clinical Practice Research Datalink (CPRD) and the Secure Anonymized Information Linkage (SAIL) found that the resulting clusters were primarily driven by seven prevalent conditions in CPRD, including depression, anxiety, allergic rhinoconjunctivitis, asthma, migraine, irritable bowel syndrome, and a range of other mental health conditions, such as obsessive-compulsive disorder, self-harm, personality disorders, and dissociative disorders [6]

The importance of considering demographic factors when examining the coexistence of multiple health conditions was emphasized by a study conducted in Fife and Tayside in Scotland which analyzed data from all registered patients aged 25 years or older using market basket analysis to identify clusters of long-term conditions. The researchers found significant variation in the patterns of condition clustering across age, sex, and socioeconomic status [24]

The presence of health conditions earlier in the sequence does not imply a causal relationship with conditions that develop later. Although associations were identified, the temporal sequence of conditions should be interpreted with caution, and further research is required to understand the mechanisms underlying these patterns.

### Strengths and limitations

This study has several strengths. It uses a high quality, representative contemporary, whole population-sample with primary care data which is a valuable source of clinical information for long-term conditions [8]. To our knowledge, it is the first study to apply this method to pregnant women. By incorporating temporal factors, this analysis overcomes the limitations of traditional clustering methods, offering a deeper understanding of the dynamic relationships and associations between different health conditions. The inclusion of these more sophisticated analytics, our approach accounts for these transitions and temporal dependencies, enabling a more accurate identification of patient trajectories. Additionally, the method employed enabled the examination of the effects of patient characteristics such as age, BMI, ethnicity, and socioeconomic status on cluster formation, providing valuable insights into how these factors influence the progression of health conditions. However, there remain some limitations. First, the true onset of a condition may be inaccurate because patients may delay visiting their GP, particularly for conditions they manage themselves such as mood disorders or those for which they get over-the-counter medications, such as allergic rhinoconjunctivitis Additionally, we did not include major conditions with lower prevalence in women of reproductive age, such as heart or renal conditions, which were less likely to appear in the clusters as a result of their small numbers. Lastly, the selection of the last pregnancy in cases where women had multiple recorded pregnancies may introduce a bias toward examining more advanced disease trajectories, particularly among older individuals.

### Implications

Understanding the order of which MLTCs are likely to emerge in an individual is important. it can help inform care pathways, support collaborative decision-making and care planning; support carers, patients, inform service development and planning, and aid in resource allocation at regional or national levels. Studying the temporal sequence of health conditions in individuals with multiple long-term conditions is a relatively new area. As such, there is relatively limited research on longer term health outcomes, particularly in younger populations and in populations like pregnant women. Studies have shown that socioeconomic deprivation and unhealthy lifestyles are associated with worse trajectories for those presenting with MLTCs, with trajectories beginning with depression being more prevalent in younger adults [25]. Therefore, the younger an individual develops a condition, the higher the likelihood of accumulating additional health issues over time, leading to more complex disease trajectories, increased healthcare needs, and higher burden for both individuals and healthcare systems [26].

In this study, the sequence clusters were driven by the first three conditions, as there was significant variability in the order of the subsequent conditions, along with low counts in later stages of disease progression. Focusing on the first three conditions allowed for a more interpretable and reliable assessment of disease progression. Owen et al in their landmark study focusing on the sequence of psychosis, diabetes and congestive heart failure argued that limiting the number of diseases or clusters in analysis will help with its interpretability and mitigate computational challenges [27]. Similarly, in this study, the number of clusters was restricted to 11 to balance granularity with interpretability. A higher number of clusters could introduce excessive complexity, making it difficult to derive meaningful insights, while too few clusters might overlook the heterogeneity in disease progression. This approach ensures that the results remain clinically relevant while maintaining computational efficiency. Owen et al’s study also demonstrated the value of studying the temporal sequence of multiple long-term conditions, as the life expectancy was different for the same conditions that developed in different orders. The choice of which 3 conditions to study was driven by data (prevalence and outcome), literature and clinical opinion. Future research on pregnant women with multiple long-term conditions may benefit from a similar approach, as limiting the number of conditions or clusters analysed can enhance methodological rigor and improve the interpretability of findings.

Cluster methods for multiple long-term conditions may have limited value for revealing important unknown groupings and risk factors because of the dominance of highly prevalent health conditions, such as mood disorders in our study sample [7]. However, their strength lies in being able to reveal previously latent/unknown associations: in our study, notably, one sequence cluster [Eating disorders or substance misuse followed by other mental health conditions and depression] identified pregnant women with multiple social and health risk factors, such as substance misuse, eating disorders, mental illness, being underweight, and social deprivation, which are all known separate associations with adverse pregnancy outcomes [28][29]. This specific combination of disorders has, to our knowledge, not been thus far recognised; yet this sequence cluster made up 6% of our study population and warrants further investigation into its outcomes.

## Conclusion

Pregnant women with pre-existing mental illness accumulate atopic conditions and other mental health conditions which contribute to multiple long-term conditions. While our findings align with existing evidence, they highlight the need for further investigation into high-risk sequences, such as those involving substance misuse and eating disorders, which may have distinct implications for pregnancy outcomes.

## Future research

This study demonstrates the utility of sequence clustering analysis to map common mental illness trajectories. Future mental health research using this technique needs to consider how to elucidate rarer sequences with high-probability transitions. One potential enhancement is reducing dimensionality by consolidating highly prevalent conditions into single variables. For instance, using a single common mental illness (CMI) variable would ensure its presence in multiple sequences while preventing it from dominating the clustering process. Similarly, conditions that often co-occur temporally, such as atopy and allergy, could be combined into a single category. This approach may help uncover different disease sequences that are currently obscured by the high prevalence of certain conditions, ultimately leading to a better understanding of mental health trajectories.

## Data availability statement

The data that support the findings of this study are available from SAIL, but restrictions apply to the availability of these data, which were used under license for the current study and so are not publicly available.

## Contributors

KN, SB, and MM conceived the study. SB and MM planned the analysis. JK prepared the data. MM undertook the analysis. SIL, KN, SB, and MM led the interpretation of results. MM and SIL drafted the manuscript. KA, HH, CMC, MU, MB, AAL, LK edited the manuscript. All authors contributed to the interpretation of the data and critically revised the manuscript for important intellectual content. All authors read and approved the final version of the manuscript and agreed to be accountable for all aspects of the work. The list of health conditions to define multimorbidity and defined the phenome were from MuM-PreDiCT consortia.

## Ethics declarations

### Ethical approval and consent to participate

I confirm that all methods were performed in accordance with the relevant guidelines and regulations, and any necessary ethics committee approvals have been obtained. All data contained in SAIL are anonymised and have permission from the relevant Caldicott Guardian or Data Protection Officer and SAIL-related projects are required to obtain Information Governance Review Panel (IGRP) approval. The SAIL Independent Information Governance Review Panel approved the study. The IGRP approval number for this study is 1287.

## Acknowledgments

This study is part of the National Centre for Population Health and Wellbeing, which is funded by Health Care Research Wales. This study makes use of anonymised data held in the Secure Anonymised Information Linkage (SAIL) Databank [8]. We would like to acknowledge all the data providers who make anonymised data available for research.

This work was funded by the Strategic Priority Fund “Tackling multimorbidity at scale” programme (grant number MR/W014432/1) delivered by the Medical Research Council and the National Institute for Health Research in partnership with the Economic and Social Research Council and in collaboration with the Engineering and Physical Sciences Research Council. BT was funded by the National Institute for Health Research (NIHR) West Midlands Applied Research Collaboration. The views expressed are those of the author and not necessarily those of the funders, the NIHR or the UK Department of Health and Social Care. The funders had no role in study design, data collection and analysis, decision to publish or preparation of the manuscript.

## Abbreviations

BMI: Body Mass Index
CMI: Common Mental Illness
EPSRC: Engineering and Physical Sciences Research Council
ICD10: International Classification of Diseases, 10th Revision
IGRP: Information Governance Review Panel
MBRRACE-UK: Mothers and Babies: Reducing Risk through Audits and Confidential Enquiries across the UK
MLTCs: Multiple Long-Term Conditions
NCCHD: National Community Child Health Dataset
NHS: National Health Service
NIHR: National Institute for Health Research
PEDW: Patient Episode Database for Wales
SAIL: Secure Anonymised Information Linkage
WDS: Welsh Demographic Service
WLGP: Welsh Longitudinal General Practice

## Supporting information

S1 File. Data quality checks

S2A-S2D Fig. Clusters revealed in the data within age, BMI, ethnicity, and deprivation level subgroups

